# Interpretable Detection of Epiretinal Membrane from Optical Coherence Tomography with Deep Neural Networks

**DOI:** 10.1101/2022.11.24.22282667

**Authors:** Murat Seçkin Ayhan, Jonas Neubauer, Mehmet Murat Uzel, Faik Gelisken, Philipp Berens

## Abstract

**Purpose:** To automatically detect epiretinal membranes (ERMs) of different sizes in various OCT scans of the central and paracentral macula region and classify them by size using deep neural networks (DNNs).

**Methods:** 11,061 OCT-images of 624 volume OCT scans (624 eyes of 461 patients) were included and graded according to the presence of an ERM and its size (small 100-1000µm, large >1000
µm). The data set was divided into training, validation and test sets (comprising of 75%, 10%, 15% of the data, respectively). An ensemble of DNNs was trained and saliency maps were generated using Guided Backprob. OCT-scans were also transformed into a one-dimensional value using t-SNE analysis.

**Results:** The DNNs’ receiver-operating-characteristics on the test set showed a high performance for no ERM, small ERM and large ERM cases (AUC: 0.99, 0.92, 0.99, respectively; 3-way accuracy: 89%), with small ERMs being the most difficult ones to detect. t-SNE analysis sorted cases by size and, in particular, revealed increased classification uncertainty at the transitions between groups. Saliency maps reliably highlighted ERMs, regardless of the presence of other OCT features (i.e. retinal thickening, intraretinal pseudocysts, epiretinal proliferation) and entities such as ERM-retinoschisis, macular pseudohole and lamellar macular hole.

**Conclusion:** DNNs can reliably detect and grade ERMs according to their size not only in the fovea but also in the paracentral region. This is also achieved in cases of hard-to-detect, small ERMs. In addition, the generated saliency maps can be used effectively to highlight small ERMs that might otherwise be missed. The proposed model could be used for screening programs or decision support systems in the future.

## 1 Introduction

Epiretinal Membrane (ERM) is a common retinal disease, occurring mainly in elderly patients, with an incidence reported between 2.2% and 28.9% [1, 2, 3]. It is characterized as an avascular fibro-cellular membrane on the innermost retinal layer and can lead to tractional changes and disruption of the retinal structure. Patients are often asymptomatic in early stages but reduced visual acuity, visual disturbances and increasing metamorphopsia are frequently seen in the later stages as the disease progresses [4]. So far, the only treatment is a vitrectomy with epiretinal peeling which increases visual acuity in most cases [5, 6].

ERMs can be identified by funduscopy or on retinal fundus images; however, the gold standard nowadays for the diagnosis is Optical Coherence Tomography (OCT), where ERMs appear as a hyperreflective membrane on the inner surface of the retina. Here, also early stages of the disease and changes of the ERM over time are visible [7]. Owing to recent advances in deep learning and the large-scale analysis of medical images via deep neural networks (DNNs) [8, 9, 10], fundus and OCT imaging modalities have been investigated for automated ERM detection from retinal images [11, 12, 13, 14, 15]. Nevertheless, these studies were limited to relatively small datasets or patients with ERM constituted only a small fraction of larger datasets [12, 14, 15, 13, 11]. In addition, these studies primarily used data from patients with advanced stages of the disease, which are comparably easy to classify. For medical reasons, it is similarly important to detect early and low-grade stages of ERMs. It is also of medical interest to segment ERMs in retinal images and some studies have automatically achieved this via DNNs trained on small data sets of 20 patients [16, 17, 18].

In this study, we investigated the automatic detection and classification of ERMs by developing an ensemble of DNNs with 11,061 clinically graded OCT-based images from 624 eyes (461 patients) and generated ensemble-based saliency maps to gain further insights into the decision-making process of the DNNs. Our model reliably detected small and large ERMs on both foveal and extrafoveal OCT scans with well-calibrated uncertainty estimates, which is unique to our study.

## 2 Methods

### 2.1 Dataset

Our dataset consisted of 624 OCT volume scans from 624 eyes of 461 patients presenting to the Department of Ophthalmology at the University of Tübingen, resulting in a total of 11,061 images. The vast majority of patients were Caucasian. All OCT images were collected with Heidelberg Spectralis OCT (Heidelberg Engineering, Heidelberg, Germany) (Table 1). The majority of the scans were standardized, horizontal fovea centered volume scans containing 25 cross-sections with a distance between the B-Scans of 61µm and a resolution of 384x496 pixels. Approximately 3% of the images were single scans through the fovea (oblique, vertical or horizontal) with a larger width and were cropped for further analysis to a standardized width of 384 pixels, focused on the fovea. An ERM was defined as a hyperreflective membrane on the inner surface ofthe retina. Eyes with secondary ERM, high myopia (<-6 D), accompanying retinal diseases, such as diabetic maculopathy, retinal vein occlusion, advanced age-related macular degeneration, previous ocular trauma or vitrectomy, and poor-quality of OCT images were excluded.

**Table 1:** Summary of the data set.

|  | Partition |  |  |
| --- | --- | --- | --- |
|  | Training (75%) | Validation (10%) | Test (15%) |
| *Number of patients* | 345 | 47 | 69 |
| *Avg. age (95% conf. int.)* | 69.25 (68.35 – 70.14) | 67.10 (64.46 – 69.74) | 69.48 (67.29 – 71.67) |
| *Number of images* | 8341 | 1146 | 1574 |
| *Laterality (L/R)* | 4174/4167 | 450/696 | 677/897 |
| *No ERM* | 3074 (36.9%) | 436 (38.0%) | 535 (34.0%) |
| *Small ERM* | 1398 (16.8%) | 204 (17.8%) | 269 (17.1%) |
| *Large ERM* | 3869 (46.3%) | 506 (44.2%) | 770 (48.9%) |

ERM: Epiretinal membrane

All images were graded by a retina specialist according to the presence of an ERM and its size, dividing them into scans without ERM, with small ERM (100-1000µm) and large ERM (>1000µm). The size of the ERM on each OCT scan was measured using a digital measurement tool and each image was classified individually. Therefore, one large membrane covering a larger area of the retina could be classified in OCT scans as small or large depending on the orientation and position of the scan. The dataset included also patients with features or entities associated with ERM, such as retinal thickening, intraretinal pseudocysts, epiretinal proliferation, ERM-retinoschisis, macular pseudohole and lamellar macular hole. To verify the dataset grading, we randomly sampled 500 B-scans representative of the three ERM classes as well as entities associated with ERM and had them re-graded by another retina specialist without disclosing the former specialist’s ERM grades. Out of 500 B-scans, the specialists disagreed on only 19 images, which led to Cohen’s kappa scores of 0.948 and 0.963 with linear and quadratic weighting schemes, respectively. A closer look into the grades of 19 B-scans revealed that the rare disagreements among graders were mostly (17 out of 19) between adjacent classes, i.e., No ERM vs Small ERM or Small ERM vs Large ERM. The graders diverged only on two B-scans by assigning No ERM or Large ERM labels in opposite scenarios. The classifications of the OCT scans were performed by two ophthalmologists who had 11 and 9 years of experience in medical retina. The study was conducted according to the guidelines and standards of the Declaration of Helsinki, and was approved by the Ethics Committee of the University of Tübingen, Germany.

### 2.2 Network Architecture and Model Development

We used the well-known ResNet50 [19] and InceptionV3 [20] architectures implemented in Keras [21] and pretrained on ImageNet [22]. We modified and fine-tuned them to our ERM classification tasks (Fig. 2). For each, we used max pooling and average pooling together at the end of the convolutional stack and concatenated their outputs. This combination had led to performance improvements in previous work [23, 24, 25, 26]. Additionally, we afterwards used two dense layers with 2048 and 512 units, which were followed by Batch Normalization [27] and ReLU activation [28]. All weight layers except the penultimate layer were equipped with *L*_2_ regularization. We employed *L*_1_ regularization to promote sparsity in the penultimate layer. Finally, we replaced the classification layer with a binary classifier with sigmoid function for basic ERM detection or a 3-way softmax classifier for detection as well as classification of ERMs according to their size.

To train and evaluate our DNNs, we used random patient-based partitions of our data (Table 1). We trained them with cross-entropy loss on the training set: 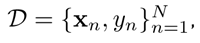 where *y_n_* is an expert-assigned label in binomial or multinomial (1-hot) representation for an image **x***_n_*. Using the multinomial representation for the sake of generality, the average cross-entropy on the training data can be expressed as follows: 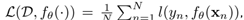 where *f_θ_*(*·*) represents a DNN parameterized by *θ*, *l*(*y_n_, f_θ_*(**x***_n_*)) =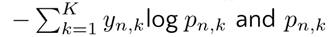 is a predicted class probability estimated via the softmax function for the *k*-th class out of *K* = 3. For *K* = 2, *l*(*y_n_, f_θ_*(**x***_n_*)) is reduced to binary cross-entropy.

We countered the class imbalance in the data with random oversampling (Table 1). Using Stochastic Gradient Descent (SGD) with Nesterov’s Accelerated Gradients (NAG) [29, 30], the minibatch size of 16, a momentum coefficient of 0.9, an initial learning rate of 0.0001, a decay rate of 0.000001 and a regularization constant of 0.00001, we trained DNNs for at least 120 epochs (see Section 2.2.1 for longer training). During the first three epochs, convolutional stacks were frozen and only dense layers were trained. Then, all layers were fine-tuned to tasks. Models with the best validation accuracy were used on the test set.

#### 2.2.1 Data augmentation and image preprocessing

To improve generalization to unseen data, we used *mixup* [32] as a data augmentation technique. Given two examples (**x***_i_, y_i_*) and (**x***_j_, y_j_*), *mixup* creates synthetic examples via their convex combinations:

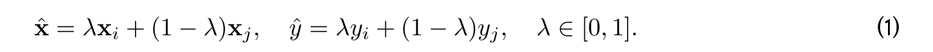

Examples were randomly drawn from training data and *λ ∼ Beta*(*α, α*) for *α ∈* (0*, ∞*). As *α →* 0, the effect of *mixup* diminishes. We used *α ∈* [0, 0.1, 0.2, 0.3, 0.4] for 120, 120, 120, 150 and 200 epochs, respectively. In the warm-up period, we set *α* = 0 for the first five epochs. In addition, we used standard data augmentation operations. The augmentation pipeline included random rotation within *±*45 degrees, horizontal and vertical translations within *±*30 pixels, brightness adjustments within *±*10%, zoom within *±*10%, and horizontal and vertical flips.

#### 2.2.2 Overconfidence and calibration of predictive probabilities

DNNs trained with hard-coded labels and cross-entropy loss are often overconfident about their predictions [33, 34, 35, 36], i.e. their predictive probabilities do not reliably indicate their expected accuracy.

Label smoothing via *mixup* can already alleviate this miscalibration of DNNs [35]. In addition to *mixup*, we used Deep Ensembles, which have been shown to improve the accuracy and calibration of DNNs [37, 38, 39]. We constructed our ensembles with five DNNs trained for ERM classification (Table 3 in Appendix), using the network architecture, hyperparameters and training procedures described above. DNNs were diversified by random initializations of dense layers, shuffling of training examples as well as mixing and data augmentation.

### 2.3 Embedding of images

To obtain insights into the feature representations learned by our ERM classification networks and their ensembles, we used t-Stochastic Neighborhood Embeddings (t-SNE) [40]. t-SNE is a non-linear dimensionality reduction method, which also allows for the interpretation of high-dimensional data in low dimensions. To evaluate ensemble-based representations, we concatenated 512 features from ensemble members’ penultimate layers and performed t-SNE based on 2560 features. We used *openTSNE* [41] with PCA initialization to better preserve the global structure of the data and improve the reproducibility [42]. We ran the optimization for 1500 iterations with a perplexity of 200, Euclidean distance and an early exaggeration coefficient of 12 for the first 500 iterations.

### 2.4 Saliency maps

Saliency maps are a post-hoc interpretability technique, often used to generate explanations for DNN decisions [43, 44, 45, 46, 47]. In the process, the prediction of a DNN is passed backwards through the network all the way back to the input image, where input pixels are associated with saliency scores according to their contribution to network outputs (Fig. 2). To compute saliency maps, we used the open-source library *iNNvestigate* [48] with the Guided Backprob algorithm [49]. This algorithm has been evaluated for clinical relevance in ophthalmology and shown to perform consistently well across different network architectures, imaging modalities, such as retinal photography and OCT, and diagnostic scenarios involving diabetic retinopathy (DR), choroidal neovascularization (CNV), diabetic macular edema (DME) or neovascular age-related macular degeneration (nAMD) [50, 51, 26].

## 3 Results

We trained DNNs to detect and classify ERMs from 11,061 individual B-scans extracted from 624 OCT volume scans. The B-scans included images with no ERMs, small ERMs and large ERMs (Fig. 1). For the binary ERM detection task, we grouped small and large ERMs together. During training, we used a recent data augmentation and regularization technique called *mixup*, along with standard data augmentation operations. The ERM detection performance of DNNs were maintained or marginally improved with the degree of mixing and longer training (Table 2. In addition to mixup, we constructed ensembles of DNNs but this also maintained the ERM detection performance across both DNN architectures, ResNet50 and InceptionV3. Given the comparable ERM detection performances of these two well-known architectures and widespread adoption of the latter for medical imaging [52, 53, 26, 54], we used InceptionV3 for both detection and classification of ERMs according to their size. Interestingly, the performances of DNNs improved with the degree of mixing and longer training in this more challenging and clinically relevant scenario (Table 3). To further improve performance, we used ensembles of DNNs once again (Table 3).

**Figure 1:**
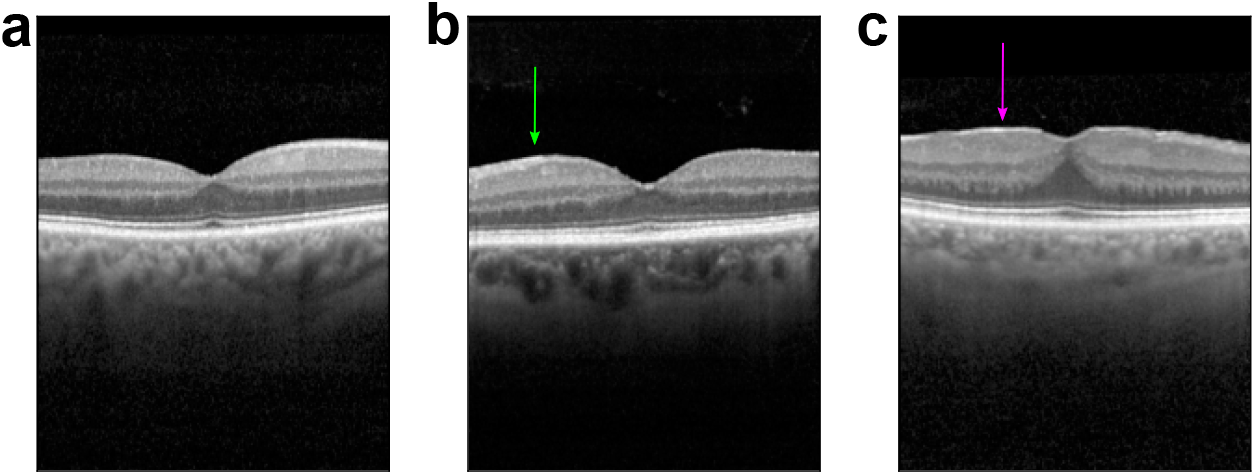
Exemplary optical coherence tomography images of the fovea. **(a)**: No epiretinal membrane (ERM). **(b)**: Small ERM (100-1000µm) (green arrow). **(c)**: Large ERM (>1000µm) (magenta arrow).

**Figure 2:**
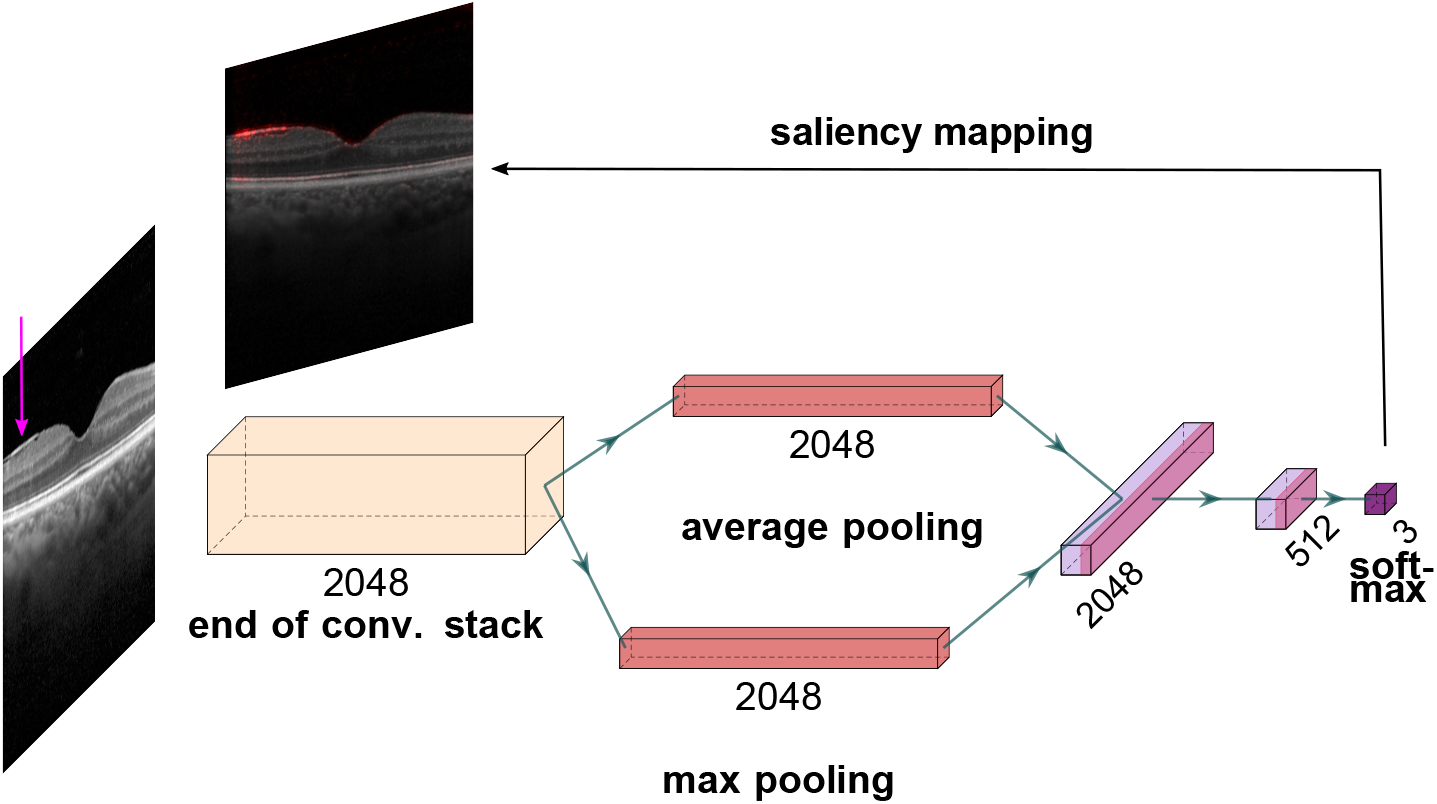
Schematic explanation of the ERM size analysis (plotted with PlotNeuralNet [31]): Given a retinal image with large ERM (indicated by an arrow in magenta), convolutional stack of the InceptionV3 architecture extracts 2048 feature maps. These are pooled with respect to average and max feature activations and fed into the fully connected (dense) layers with 2048 and 512 units. Finally, a 3-way softmax function achieves classification based on 512 features from the penultimate layer and detects large ERM. A saliency map highlights the regions relevant to the DNN decision.

**Table 2:**
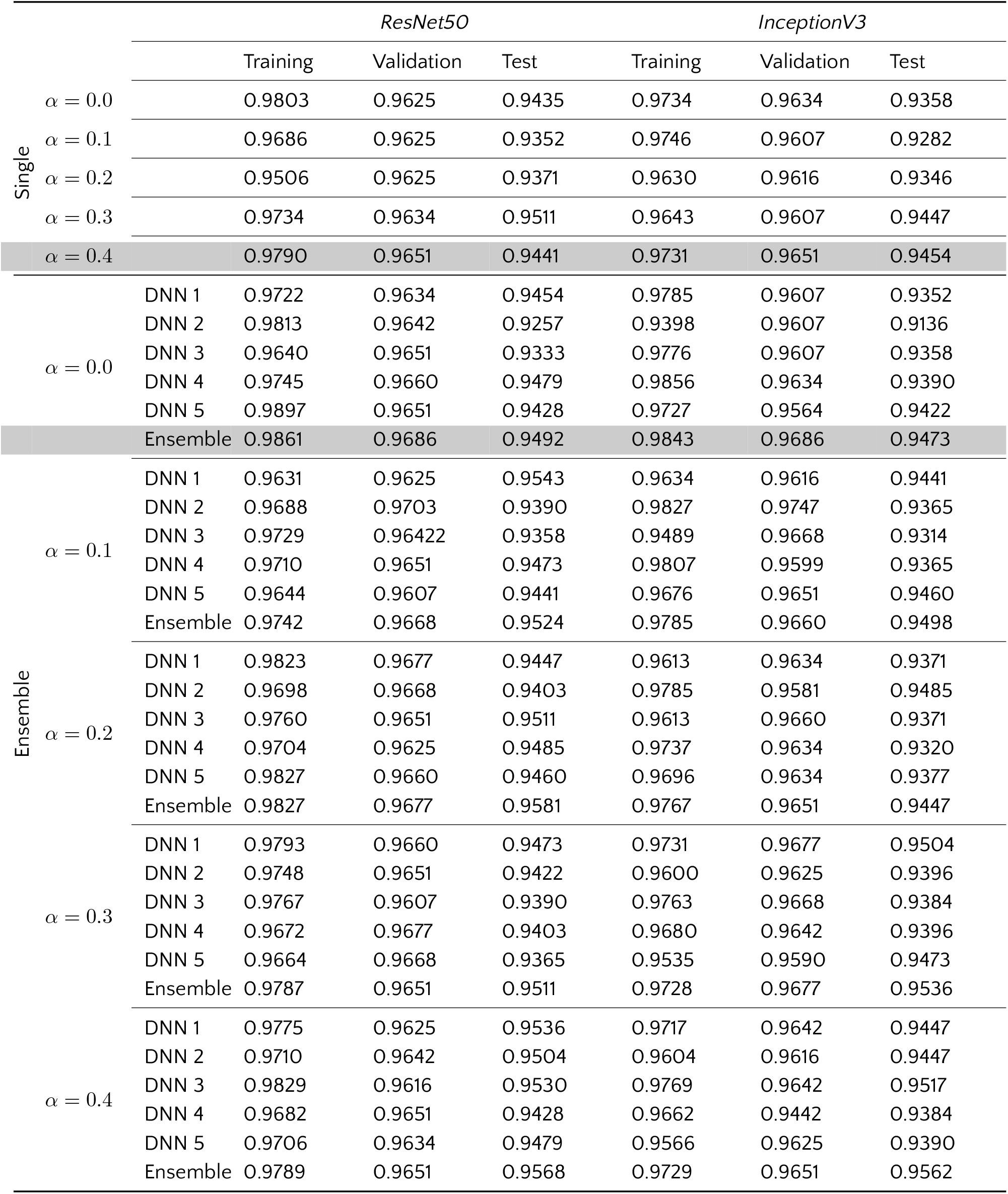
ERM detection accuracy of individual networks and their ensembles for various degrees of mixing (indicated by *α*) and corresponding numbers of epochs as given in Section 2.2.1. Gray rows indicate the models of choice based on validation accuracy.

Our 3-way classification DNNs accurately detected the presence of ERM in retinal images and were also able to determine the ERM size, which is an important feature of clinical relevance, with high accuracy (Table 3, Fig. 3). The best ensemble model was obtained from DNNs trained with intense mixing (*α* = 0.4) for 200 epochs, achieving a 3-way classification accuracy of 89.33%) (indicated by the gray row in Table 3) and an AUC of 0.99, 0.92 and 0.99 for normal B-scans, B-scans with small and large ERMs, respectively. Interestingly, small ERMs were more difficult for DNNs to detect than the large ones (Fig. 3 (a) and (b)). While this difficulty can be attributed to the ERM pathophysiology that causes small ERMs to be the most difficult to identify among the three classes, ablation of ensembling along with mixup led to inferior performance and impacted the small ERM detection rate most (see supplementary Fig. 7 (a) and (b)).

**Table 3:**
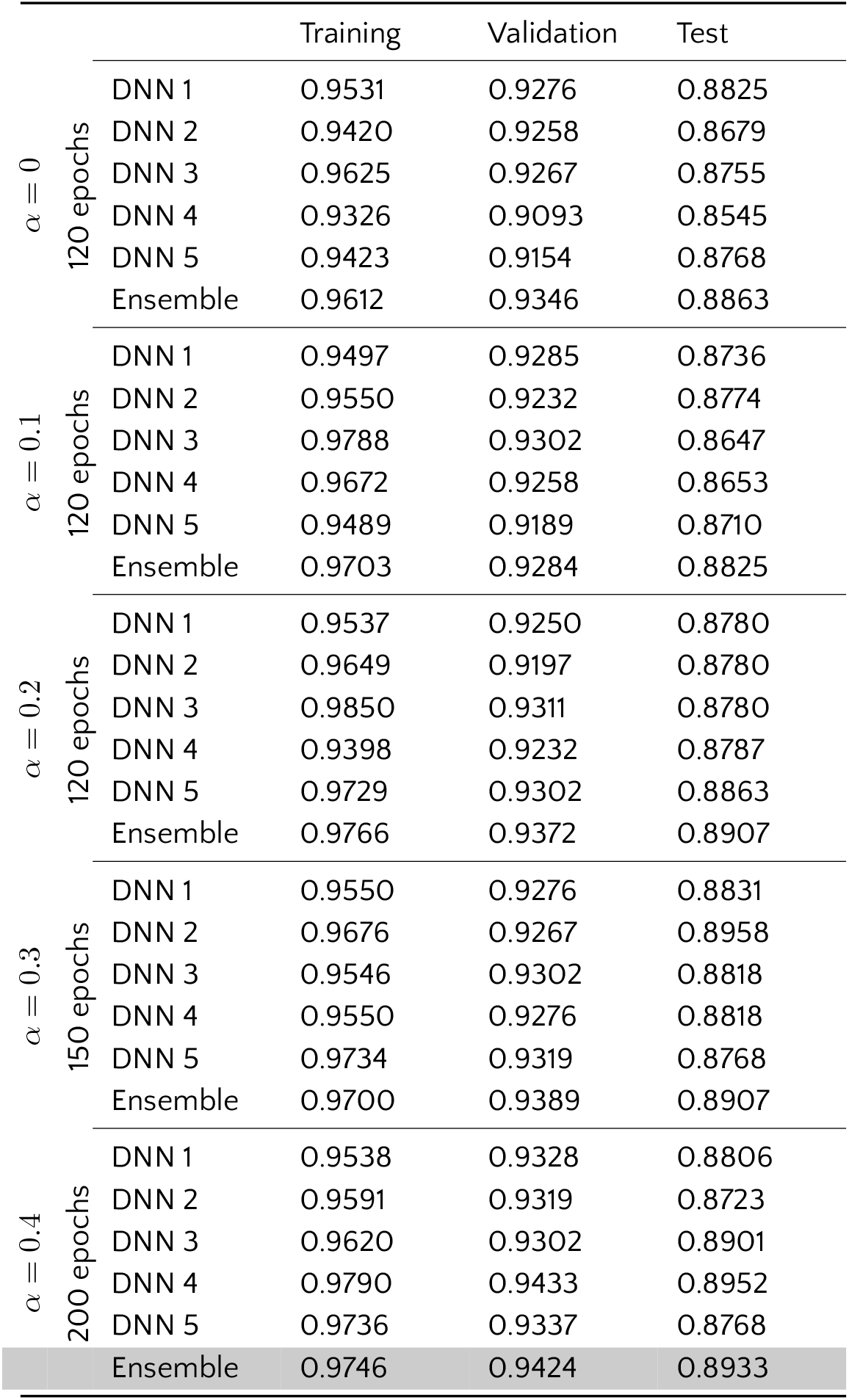
ERM classification accuracy for individual networks and their ensembles for various degrees of mixing (indicated by *α*) and numbers of epochs. Gray row indicates the ensemble of choice for further analysis.

**Figure 3:**
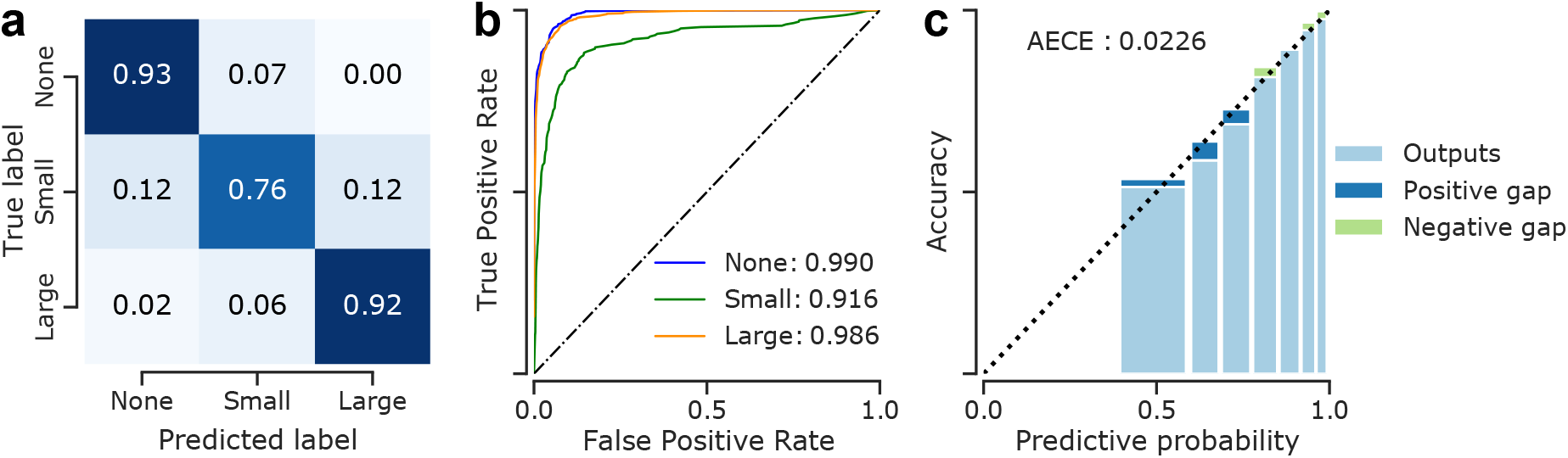
Detailed analysis of classification performance on the test set. **(a)**: Confusion matrix for the selected ensemble model. **(b)**: Receiver operating characteristics of the ensemble model. Numbers indicate AUC scores for the respective classes. **(c)**: Reliability diagrams and calibration of the selected ensemble. The calibration error was estimated via reliability diagrams [55, 33, 56] and the Adaptive Expected Calibration Error (AECE) [55]. While a negative gap (light green) between predictive probability and accuracy indicates a lack of confidence in predictions, a positive gap (dark blue) points at overconfidence.

We also assessed the uncertainty estimates provided by our ensemble model and found that its predictive probabilities for the training and validation data were slightly oversmoothed, likely due to the combined effects of *mixup* and ensembling, but overall well-calibrated on the test data with a small adaptive expected calibration error of 0.02 (Fig. 3 c). With the ablation of ensembling and mixup, the calibration error was 0.05 (Fig. 7 c). This indicated that the uncertainty estimates reported by our ensemble with mixup can be expected to provide useful information about the performance of the model, with high uncertainties corresponding to more errors, an information which may be taken into account by clinicians making decisions based on the DNNs outputs.

To better understand which areas of the images were important to the DNN’s decision-making process, we created saliency maps using the Guided Backprob algorithm [49]. These saliency maps clearly highlighted important areas of interest mainly on the inner surface of the retina (Fig. 4 d-l), including scans with a small ERM. This is remarkable, since many of the small ERMs are very fine structures which are difficult to detect even for ophthalmologists. It should also be emphasized that associated retinal changes and entities (i.e. intraretinal pseudocysts, retinoschisis, lamellar macular hole) were not markedly highlighted on the saliency maps and thus seemed to be not crucial for the decision-making process of the DNN. In the cases of images without ERM, vitreous opacities were frequently marked (Fig. 4 a-c).

**Figure 4:**
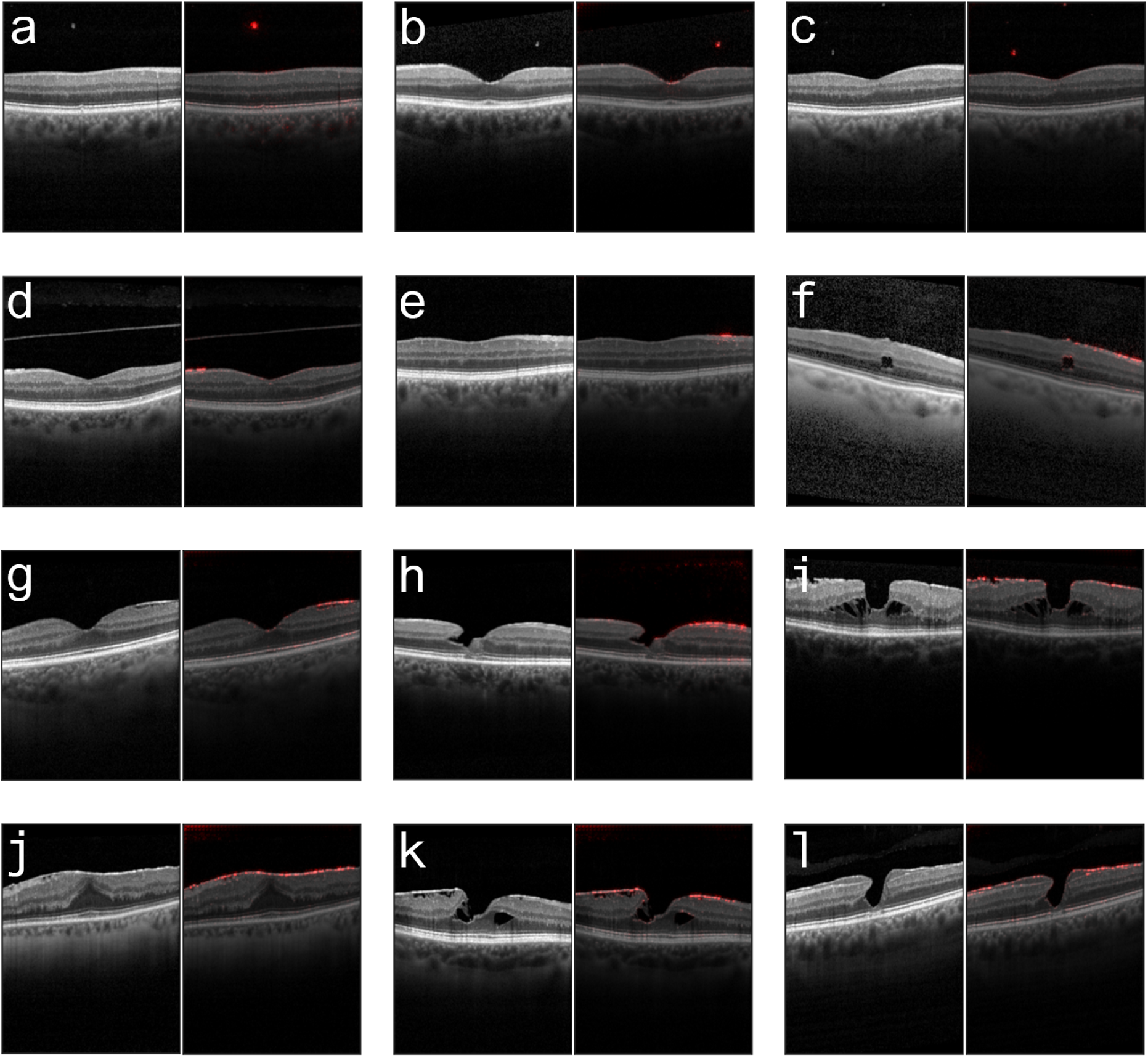
Examples of the correctly classified optical coherence tomography (OCT) images (left) and generated saliency maps (right), showing reliable highlighting of ERM in foveal and extra-foveal areas. **(a-c)**: OCT scans without ERM and highlighted opacities in the vitreous. **(d)**: Foveal scan with a small extrafoveal ERM. **(e)**: Extrafoveal scan with a small ERM. **(f)**: Extrafoveal scan with a large ERM and an intraretinal pseudocyst. **(g)**: Foveal scan with a large extrafoveal ERM not affecting the foveal depression. **(h)**: Lamellar macular hole with a large ERM. **(i)**: A large ERM and foveoschisis. **(j)**: Foveal scan with a large ERM and a elevated foveal depression. **(k)**: Foveoschisis with a large ERM. **(l)**: Early stage of a lamellar macular hole with a large ERM and epiretinal proliferation.

The proposed model classified 169 images of the test set (1574 images) differently than the retina specialist (Fig. 5). Re-analysis of these images by two additional retina specialists revealed that 53 scans had indeed been correctly classified by the DNN and that most of these grader misclassifications were due to obvious documentation errors (for example, 24 of the 53 images originated from one volume scan of one patient with prominent ERM and were graded as “small ERM” by the human). In the clinical context, however, it is far more interesting to see how often the algorithm missed an ERM, which was the case in 62 scans. Upon inspection, we found that these were mostly very fine and small ERMs at the edge of what was considered as ERM present (> 100µm) (Fig. 5a).

**Figure 5:**
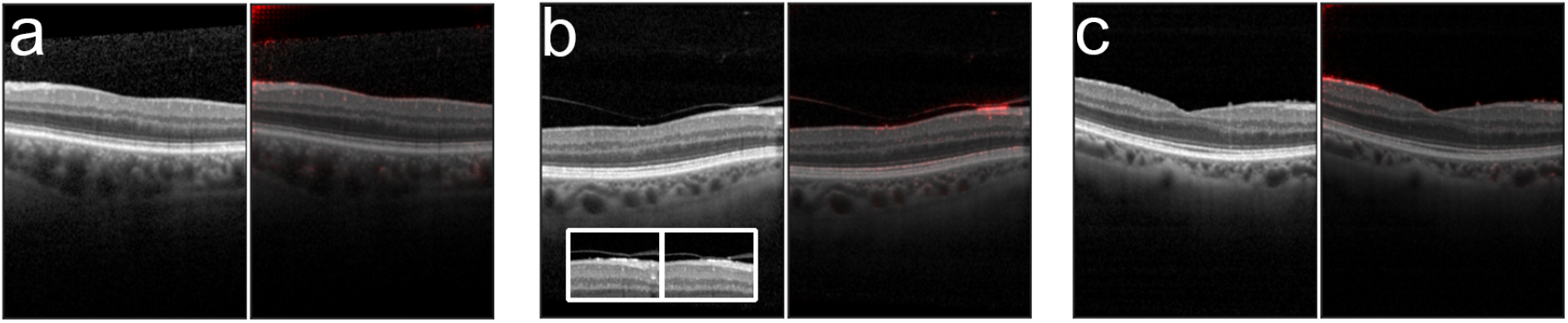
Examples of incorrectly classified optical coherence tomography (OCT) scans (left) and generated saliency maps (right). **(a)**: This scan was classified as “no ERM”, while a small ERM of about 170µm size is detectable. **(b)**: Even though the ERM is highlighted in the saliency map the model classified this image incorrectly. The neighbouring OCT images superior and inferior (presented in the box) to this one were classified correctly. **(c)**: A large ERM that has been classified by the model as a small ERM.

To study the representation learned by the DNNs, we embedded all images in our dataset into one dimension using t-Stochastic Neighborhood Embedding (t-SNE) based on the network activations (features) in the penultimate layers of the ensemble members (Fig. 6, also see Methods). In this representation, each dot corresponds to a single OCT B-scan, with the color indicating its class. We found that the resulting t-SNE map (Fig. 6a) followed the disease continuum with great accuracy ordering the discrete classes accordingly with ERM-negative cases being placed to the left and positive ones towards right. Pairing the t-SNE coordinates with predictive uncertainty associated with retinal images, we also found that the average uncertainty was highest at the boundaries between the stages of ERM (Fig. 6a). High uncertainty was also indicative of wrong predictions (Fig. 6b), in line with the finding that most misclassifications occurred at the transition between ERM-negative and small ERMs, and the good calibration of the model. In fact, incorrect and correct predictions were coupled with significantly higher and lower uncertainty, respectively (*p <* 0.0001, Mann-Whitney U test, *n_wrong_*= 446*, n_correct_* = 10, 615) (Fig. 6c).

**Figure 6:**
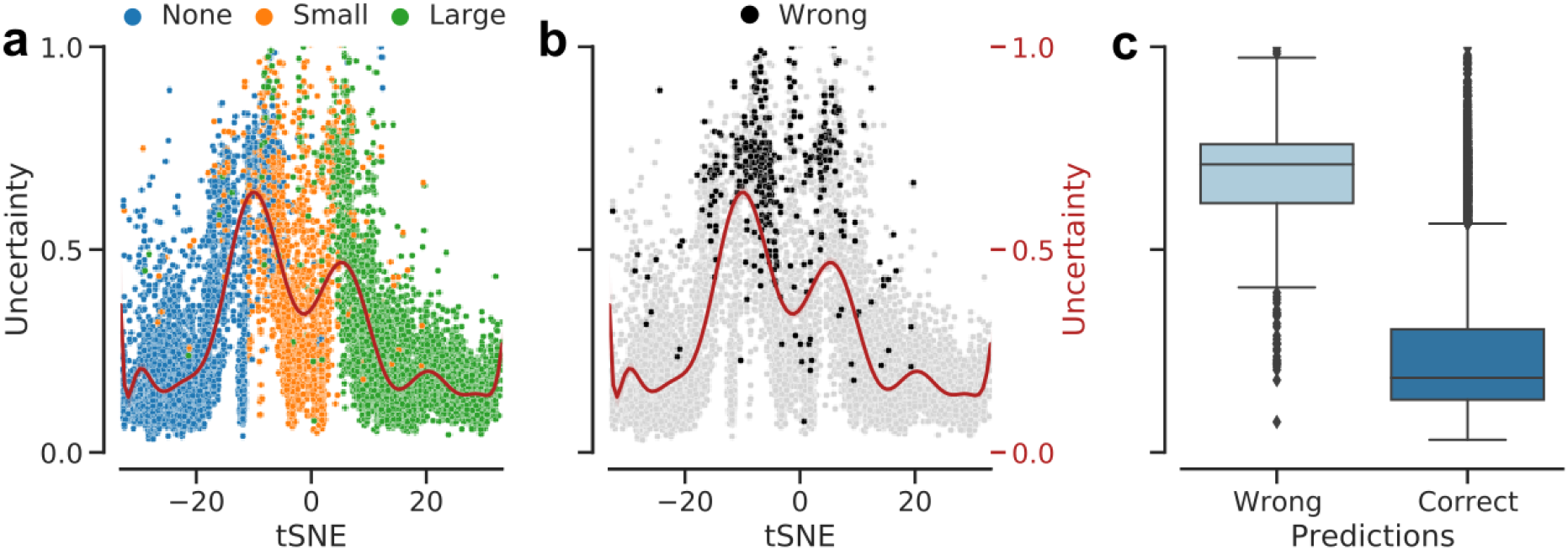
1D t-SNE map via the best ensemble model and predictive uncertainty difference between the groups of wrong and correct predictions. The paired t-SNE coordinates with predictive uncertainty measured by the entropy of the ensemble output and fitted a polynomial regression curve to data. **(a)**: Training, validation and test data aligned together and colored with respect to the ERM labels. **(b)**: Same map as in (a) but colored w.r.t. correct (gray) and wrong (black) predictions. **(c)**: Uncertainty distribution in groups of wrong and correct predictions.

**Figure 7:**
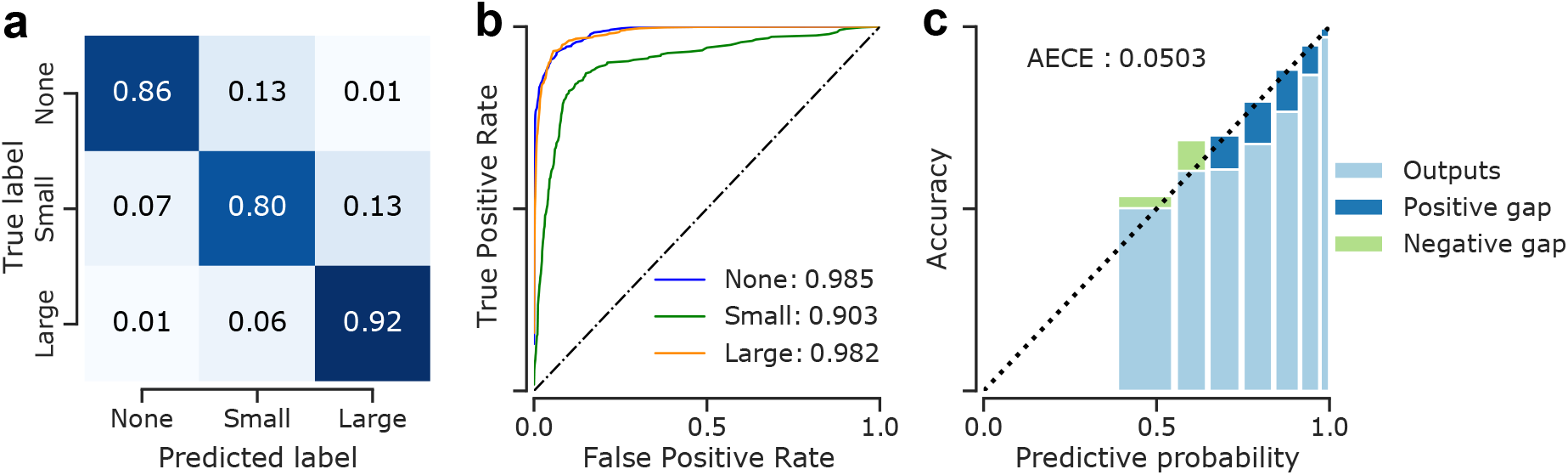
Detailed analysis of classification performance of “DNN 1” from the ensemble members trained without *mixup* (Table 3) on the test set. **(a)**: Confusion matrix for the model. **(b)**: Receiver operating characteristics of the model. Numbers indicate AUC scores for the respective classes. **(c)**: Reliability diagrams and calibration of the model.

## 4 Discussion

We showed that DNNs can reliably detect ERMs of different severity on OCT images of the macula, classify them based on their size and provide well-calibrated uncertainty estimates for their decisions. In addition, to gain further insights into the DNNs’ inner workings leading to ERM decisions, we computed ensemblebased saliency maps [26] that labeled ERMs independently of accompanying retinal changes with high confidence.

As the OCT exam provides a detailed visualization of the posterior pole and the retinal layers, various ERM classification systems have been proposed. Some distinguish between attached and detached vitreous, central retinal thickness or classify patients based on fovea involvement and changes in retinal layers [57, 58, 4]. However, these grading systems are only applicable to OCT images of the fovea and have not gained general acceptance in the clinical routine. As the aim of our work was to provide an automatic system for robust detection of ERM, we therefore included not only single fovea scans but also OCT images from the paraand perifoveal region. In order to classify images with ERM further, we decided to group them by size as larger ERMs tend to alter retinal layers more severely than smaller ERMs. We developed an ensemble of DNNs to automatically detect and grade ERMs on foveal and extrafoveal OCT scans and our proposed model showed high accuracy in this multi-class classification scenario. In this respect, our study makes progress in follow-up to the existing work on ERM detection from retinal images, which so far considered binary scenarios, i.e., deciding either the absence or presence of ERM [11, 12, 13, 14, 15]. In fact, our proposed DNN seems to grade the extend of the disease automatically. Despite being presented only categorical labels and having no explicit knowledge of the ERM pathology or development, the generated one dimensional t-SNE map (Fig. 6a) indicates that the model faithfully restored the disease continuum from data and ordered the classes accordingly with ERM-negative cases being placed to the left, small ERMs in the center and large ERMs towards the right. Similar characteristics have been observed in the past for automatic ordering of diabetic retinopathy stages [25].

Our DNNs misclassified only a few large ERM scans as healthy, demonstrating that a more pronounced disease is easier to recognize on OCT scans. Furthermore, it should be noted that these misclassified scans presented only as a very fine hyperreflective line and could have been easily missed even by ophthalmologists. As one would therefore expect, small ERMs were more difficult for our DNNs to detect, compared to the large ones or cases of no ERM. Belonging to a transitional state between no ERM and advanced stages, the examples of small ERM also demonstrated an overlap with the other two classes. The predictive uncertainty of the proposed model was indicative of such difficult cases mostly located around decision boundaries. Interestingly, the peak uncertainty was observed when transitioning from ERM-negative to small ERM scans, indicating the difficulty of detecting such early-onset cases, similar to the mild DR cases reported previously [25]. Distinguishing between adjacent stages seems to be not only for DNNs but also for retina specialists challenging, as the vast majority of the few disagreements among the two graders were between adjacent stages.

The accuracy claims of other studies have to be interpreted therefore in light of the data used to train and evaluate their models, typically omitting small ERMs. Previous publications have often not provided much information on the data used, with regards to the severity of the ERM or OCT characteristics, but the shown examples indicate more advanced cases. Only Parra-Mora et al. [15] stated that they used OCT scans of different disease severity, without further specifying the distribution of the stages. In contrast, the distribution of ERMs in the study presented here allows for a more accurate representation in terms of disease severity of the study population, as we collected a large dataset derived from the clinical routine of our clinic. Consequently, we covered the whole spectrum of different ERMs presenting with features (i.e. retinal thickening, intrartinal pseudocysts, retinoschisis) and other entities (i.e. ERM-retinoschisis, lamellar macular hole, macular pseudohole) associated with ERM. We implemented not only standardized horizontal OCT scans but also vertical and oblique oriented ones. In addition to treating ERM detection as a classification task as in this study, segmentation algorithms have also been trained to detect ERMs [16, 17, 18]. Some of these works have primarily focused on the detection of the internal limiting membrane (ILM), which might be erroneous in eyes with a visible vitreous or posterior vitreous limiting membrane - as regularly seen in patients with ERM (i.e. Fig. 4 d, Fig. 5 b).

Our study presents a path for a robust detection of ERMs in a variety of patients, with different accompanying features and with different scan patterns. Also, explanations of our DNNs using saliency maps have highlighted the usefulness of the model in detecting ERMs, even if they are small and not located in the fovea. Vitreous opacities were frequently marked in images without ERM (i.e. Fig. 4 a-c). This can be interpreted in accordance with the fact that saliency maps often appear diffuse and not well localized in the absence of pathologies which are supposed to be detected by the DNN. Previous ERM classification studies have relied on the GradCAM algorithm [59] to generate saliency maps which highlighted the approximate region of the retina where the ERM was detected [12, 15]. In contrast, our proposed saliency maps based on GuidedBackProp [49, 26] presents substantially more detail and a much finer resolution, while reliably detecting the ERM at the same time. Interestingly, saliency maps repeatedly marked outer retinal layers like the retinal pigment epithelium under the ERMs as important areas (Fig. 4). A possible explanation for this could be that the ERM casts a light shadow over the underlying retinal structures, which could be detected by the model and used as an indirect OCT retinal biomarker. The precise representation of ERMs in the presented saliency maps could help to increase trust in the described DNN as it makes the decision-making process of the algorithm more understandable for patients and physicians. To this end, visual counterfactual explanations (VCEs) via adversarially robust ensembles [60] can be also used to augment saliency maps with more realistic and reliable visualizations in future studies.

Due to the increasing age of the general population it will be challenging to provide comprehensive ophthalmological care in the near future, thus shifting the focus to decision support systems or automatic screening approaches [61]. This is particularly the case for ERMs, since it usually starts as a monocular disease that is not always directly noticed by the patient and is probably associated with a worse visual prognosis in case of delayed treatment [62]. At the same time, the amount of data generated per patient and visit increases over the years, making it more difficult for a physician to accurately assess all of it. Thus, our state-of-the-art DNN could help ophthalmologists in the sense of a decision-support system and, for instance, point out abnormal areas of volume scans in order to prevent medical misdiagnosis and improve the quality of healthcare. Multi-task models are also promising for more comprehensive systems that will take into account multiple pathologies associated with ERM, as recently demonstrated for the case of nAMD activity detection [54].

A limitation of the current study, however, can be thought of as the position and orientation of OCT scans, which could have led to different ERM size measurements under different settings. To address this, an alternative would be to display the entire ERM on one OCT volume scan, segment it and finally measure the largest diameter or the area covered by the ERM. However, this is, due to technical reasons, in many cases not possible and it would be practically impossible to obtain these labels for such a large data set in a clinical setting. A possible avenue to explore in this regard is the use of self-supervised learning and *foundation models* [63] that would benefit from broad data and sidestep the need for expert labels.

## Data Availability

The optical coherence tomography scans were obtained from the University Eye Clinic and their use was permitted by the Institutional Ethics Committee of the University of Tuebingen.

## Acknowledgments

We thank the German Ministry of Science and Education (BMBF) for funding through the Tübingen AI Center (FKZ 01IS18039A) and the German Science Foundation for funding through a Heisenberg Professorship (BE5601/4-2) and the Excellence Cluster “Machine Learning — New Perspectives for Science” (EXC 2064, project number 390727645). MMU received a postdoctorate international research grant (grant number: BIDEB-2219) from “The Scientific and Technological Research Council of Turkey—TUBITAK.” The funding bodies did not have any influence in the study planning and design.

## Appendix

A.1 Classification performance

A.2 Ablation of ensembling and mixup

## Notes

### Competing Interest Statement

The authors have declared no competing interest.

### Funding Statement

We thank the German Ministry of Science and Education (BMBF) for funding through the Tuebingen AI Center (FKZ 01IS18039A) and the German Science Foundation for funding through a Heisenberg Professorship (BE5601/4-2) and the Excellence Cluster "Machine Learning - New Perspectives for Science" (EXC 2064, project number 390727645). MMU received a postdoctorate international research grant (grant number: BIDEB-2219) from "The Scientific and Technological Research Council of Turkey-TUBITAK." The funding bodies did not have any influence in the study planning and design.

### Author Declarations

Our dataset consisted of 624 OCT volume scans from 624 eyes of 461 patients presenting to the Department of Ophthalmology at the University of Tuebingen. The study was conducted according to the guidelines and standards of the Declaration of Helsinki, and was approved by the Ethics Committee of the University of Tuebingen, Germany.

### Summary of Updates

We updated the manuscript as a result of the second round of revisions. Changes were made to clarify a few things about expert-labeling of ERMs via a digital tool as well as the experience of clinicians. Also, we added a paragraph in Discussion to reflect on the limitations of the study.

